# Racial and Ethnic Disparities in Access to Health Care Among Adults in the United States: A 20-Year National Health Interview Survey Analysis, 1999–2018

**DOI:** 10.1101/2020.10.30.20223420

**Authors:** César Caraballo, Dorothy Massey, Shiwani Mahajan, Yuan Lu, Amarnath R. Annapureddy, Brita Roy, Carley Riley, Karthik Murugiah, Javier Valero-Elizondo, Oyere Onuma, Marcella Nunez-Smith, Howard P. Forman, Khurram Nasir, Jeph Herrin, Harlan M. Krumholz

## Abstract

**Importance:** Racial and ethnic disparities plague the US health care system despite efforts to eliminate them. To understand what has been achieved amid these efforts, a comprehensive study from the population perspective is needed.

**Objectives:** To determine trends in rates and racial/ethnic disparities of key access to care measures among adults in the US in the last two decades.

**Design:** Cross-sectional.

**Setting:** Data from the National Health Interview Survey, 1999–2018.

**Participants:** Individuals >18 years old.

**Exposure:** Race and ethnicity: non-Hispanic Black, non-Hispanic Asian, non-Hispanic White, Hispanic.

**Main outcome and measures:** Rates of lack of insurance coverage, lack of a usual source of care, and foregone/delayed medical care due to cost. We also estimated the gap between non-Hispanic White and the other subgroups for these outcomes.

**Results:** We included 596,355 adults, of which 69.7% identified as White, 11.8% as Black, 4.7% as Asian, and 13.8% as Hispanic. The proportion uninsured and the rates of lacking a usual source of care remained stable across all 4 race/ethnicity subgroups up to 2009, while rates of foregone/delayed medical care due to cost increased. Between 2010 and 2015, the percentage of uninsured diminished for all, with the steepest reduction among Hispanics (−2.1% per year). In the same period, rates of no usual source of care declined only among Hispanics (−1.2% per year) while rates of foregone/delayed medical care due to cost decreased for all. No substantial changes were observed from 2016–2018 in any outcome across subgroups. Compared with 1999, in 2018 the rates of foregone/delayed medical care due to cost were higher for all (+3.1% among Whites, +3.1% among Blacks, +0.5% among Asians, and +2.2% among Hispanics) without significant change in gaps; rates of no usual source of care were not significantly different among Whites or Blacks but were lower among Hispanics (−4.9%) and Asians (−6.4%).

**Conclusions and Relevance:** Insurance coverage increased for all, but millions of individuals remained uninsured or underinsured with increasing rates of unmet medical needs due to cost. Those identifying as non-Hispanic Black and Hispanic continue to experience more barriers to health care services compared with non-Hispanic White individuals.

**KEY POINTS:** *Question:* In the last 2 decades, what has been achieved in reducing barriers to access to care and race/ethnicity-associated disparities?

*Findings:* Using National Health Interview Survey data from 1999–2018, we found that insurance coverage increased across all 4 major race/ethnicity groups. However, rates of unmet medical needs due to cost increased without reducing the respective racial/ethnic disparities, and little-to-no change occurred in rates of individuals who have no usual source of care.

*Meaning:* Despite increased coverage, millions of Americans continued to experience barriers to access to care, which were disproportionately more prevalent among those identifying as Black or Hispanic.

## BACKGROUND

The United States (US) health system has been plagued by racial and ethnic disparities in access to health care.^1-3^ Hispanic and Black individuals experience greater barriers to health care services, such as lack of health insurance coverage, lack of a usual source of care, and unmet medical needs due to cost.^4-8^ Thirty-five years ago, in 1985, the US Department of Health and Human Services published a landmark report, Black and Minority Health,^9^ which highlighted racial and ethnic health disparities. In the report, the Secretary’s Task Force recommended increasing minorities’ health insurance coverage to facilitate their access to health care. Subsequently, in 2003, the Institute of Medicine’s report, Unequal Treatment, highlighted again how minorities continued to experience greater barriers to health care services.^10^ Since these reports were published, major national public health policies and programs have been implemented to address racial and ethnic disparities in access to health care, such as the Healthy People Initiative and the Affordable Care Act (ACA).^11-13^ Amid these efforts to address disparities in access to care, it is important to assess what has been achieved over the last 20 years.

While a number of studies have reported on the status of racial and ethnic health care disparities in the US,^1-8^ they were limited by short study periods—most are single-year studies— or focused on the years surrounding the passage of the ACA. We lack a multi-decade, comprehensive view of the racial and ethnic trends in health care access in the US.

The National Health Interview Survey (NHIS) is the principal federal source of health information on the civilian, non-institutionalized population of the US and provides an ideal platform to evaluate racial and ethnic differences in health care access over the past 2 decades. Accordingly, we used the NHIS to evaluate racial and ethnic differences in the various facets of access to care barriers, using key indicators of health care accessibility and affordability such as lack of insurance coverage, lack of a usual source of care, and foregone or delayed medical care due to cost—some of the most commonly used metrics in access to health care studies.

## METHODS

### Data Source

We used data from the annual cross-sectional surveys of the NHIS for the years 1999 to 2018. The survey uses a complex multistage area probability design that accounts for non-response and allows for nationally representative estimates, including underrepresented groups.^14^ We obtained the data from the Integrated Public Use Microdata Series (IPUMS) Health Surveys, which is a hub that provides harmonized data for extraction and analysis.^15^ The code used to analyze these data are available from the corresponding author upon reasonable request. This study received exemption for review from the Institutional Review Board at Yale University as NHIS data are publicly available and deidentified.^16^

### Study Population

We included all individuals >18 years old from years 1999 to 2018 from the Sample Adult Core file, which contains the responses from one adult who is randomly selected from each family for a more in-depth questionnaire. We excluded those with unknown race or ethnicity information. Due to limited sample size, we did not include those not from Hispanic ethnicity who identified their primary race as Alaskan Native or American Indian, Other Race, or who did not select a primary race.

### Study Outcomes

The main outcomes of the study were: lack of health insurance coverage, foregoing and/or delaying medical care due to cost, and not having a usual source of care. Individuals were classified as “uninsured” if at the time of interview they reported not having any private health insurance, Medicare, Medicaid, military plan, government- or state-sponsored health plan, or if they had only Indian Health Service coverage.^17^ Consistent with Data Briefs from the National Center for Health Statistics^18^ and prior NHIS studies,^19-23^ we used survey responses that indicated foregone or delayed medical care due to cost and not having a usual source of care as additional indicators of barriers to health care access. Foregone or delayed medical care was defined as answering “Yes” to any of the following three questions: (during the past 12 months) “Has medical care been delayed for you because of worry about the cost?”; “Was there any time when you needed medical care, but did not get it because you couldn’t afford it?”; or “Was there any time when you needed prescription medicines but didn’t get it because you couldn’t afford it?.” Not having a usual source of care was defined as answering “No” to the question “Is there a place that you usually go to when you are sick or need advice about your health?” For each of these 3 outcomes and each study year, we calculated racial/ethnic “disparities” as the absolute prevalence gap between non-Hispanic White and the other race/ethnicity subgroups defined below.

### Variables of Interest

#### Race and Ethnicity

Hispanic ethnicity was defined as answering “Yes” to the question, “Do you consider yourself Hispanic/Latino?” Race was ascertained by the response to the questions, “What race do you consider yourself to be?” and, if more than one was reported, “Which one of these groups would you say BEST represents your race?” Race was then categorized as White, Black/African-American, or Asian (which included Chinese, Filipino, Asian Indian, and Other Asian). We created 4 mutually exclusive subgroups based on primary race selected and ethnicity combination: non-Hispanic White, non-Hispanic Black, non-Hispanic Asian, and Hispanic. For simplicity, we hereafter refer to these groups as White, Black, Asian, and Hispanic.

#### Respondent Characteristics

Covariates of interest were age in years, and region (Northeast, North Central/Midwest, South, West). Other descriptive variables were sex, education level (less than high school, high school diploma or GED, some college [including associate degree], Bachelor’s degree or higher), US citizenship status, marital status (married/living with partner vs. not), employment status (working, not in labor force, unemployed), comorbidities (hypertension, diabetes, prior stroke or myocardial infarction, cancer, emphysema, or chronic bronchitis), smoking status, obesity, and flu vaccination in the past 12 months. We also included annual family income based on percent of family income relative to the respective year’s federal poverty limit from the US Census Bureau^24^ using categories consistent with prior NHIS studies: middle/high income (≥200% federal poverty limit) or low income (<200% federal poverty limit).^19,25^

### Statistical Analysis

All analyses used methods appropriate for structured survey data, incorporating strata and weights to produce nationally representative estimates. All person weights were pooled and divided by the number of years studied, in accordance with NHIS guidance.^26^ We first summarized all respondent characteristics by race/ethnicity. We then estimated the annual outcome rates for each race/ethnicity subgroup using a linear probability model, with the outcome as the dependent variable, age (discrete), a dummy variable for each region (Northeast, North Central/Midwest, South, West) as independent variables, and an indicator for each year of interview. Age and region indicators were centered on their overall mean for the study sample and the intercept was suppressed; the coefficients for each year then represented age- and region-adjusted annual outcome rates. Then, from the annual prevalence of each outcome among those identifying as Black, Asian, or Hispanic, we subtracted the annual prevalence of the same outcome for the same year among those identifying as White (e.g. proportion of Hispanic people uninsured in 2010 - proportion of White people uninsured in 2010). We used this gap as our indicator of racial/ethnic disparities using those identifying as White as the reference group. Using these annual rates, we then estimated trends in outcomes rates and disparities by fitting regression models for each race/ethnicity subgroup. The dependent variable was the age- and region-adjusted annual prevalence of each outcome or disparity (calculated as described earlier), and the independent variable was time in years. Rather than assume a monotone relationship between time and outcome rates, we graphically assessed the relationship first; based on this assessment, we modelled time as a linear spline with knots at 2010 and 2016 to reflect observed inflection points. To account for serial correlation of annual outcome rates, we incorporated an autoregressive error term with 1-year correlation. We then used the coefficient of the time variables to evaluate the slope of each outcome’s prevalence during each period. In a separate analysis, we estimated the absolute difference in prevalence of each outcome and disparity between 1999 and 2018 by incorporating the standard error of each estimate to account for its precision. “Don’t know,” “Refused,” or no response values were set to missing for each outcome. For all analyses, two-sided P-value <0.05 was used to determine statistical significance. All analyses were performed using Stata SE version 16.1 (StataCorp, College Station, TX).

## RESULTS

Of 603,140 adults interviewed from 1999 to 2018, we excluded 112 with missing race/ethnicity information and 6,673 who identified as non-Hispanic Other Race (which included non-Hispanic Alaskan Native or American Indian, Other Race, or no primary race selected) from our analysis. Thus, our study sample included 596,355 adults (**Supplementary Figure S1**). Of these, 69.7% (weighted proportion; 95% CI: 69.3%, 70.2%) identified as White, 11.8% (95% CI: 11.5%, 12.1%) identified as Black, 4.7% (95% CI: 4.6%, 4.8%) identified as Asian, and 13.8% (95% CI: 13.5%, 14.2%) identified as Hispanic. In addition, to illustrate how the subgroups may have changed during the study period, general characteristics of the study population by race/ethnicity in the first 2 years (1999 and 2000) and last 2 years (2017 and 2018) of interview are shown in **Supplementary Table S1** and the annual distribution of race/ethnicity over the 20-year study period is shown in **Supplementary Figure S2**.

### Health Insurance Coverage

The age- and region-adjusted annual percentages of uninsured individuals by race/ethnicity are shown in **Figure 1**, and the annualized trends by study periods are shown in **Table 1**. The overall missing rate of insurance status was 0.44%. Before 2010, prevalence trends were stable for all 4 race/ethnicity subgroups and were without significant changes in disparities except for those identifying as Asian, for whom the gap with those identifying as White had a slightly negative trend (−0.3% per year, 95% CI: −0.7%, −0.1%). Between 2010 and 2015, the percentage of uninsured diminished at an annual rate of −0.89% (95% CI: −1.25%, −0.05%) for those identifying as White, −1.34% (95% CI: −2.20%, −0.48%) for those identifying as Black, - 1.23% (95% CI: −2.0%, −0.45%) for those identifying as Asian, and −2.12% (95% CI: −3.05%, - 1.21%) for those identifying as Hispanic. During this period, the gap in uninsured rates narrowed between those identifying as White and those identifying as Hispanic (−1.2% per year, 95% CI: - 1.8%, −0.6%). No significant trends in overall prevalence or disparities were observed between 2016 and 2018 across subgroups. The absolute change in percentage of uninsured between 1999 and 2018 was −2.9% (95% CI: −3.6%, −2.2%) for those identifying as White, −6.4% (95% CI: - 8.7%, −4.1%) for those identifying as Black, −9.5% (95% CI: −13.3%, −5.7%) for those identifying as Asian, and −9.0% (95% CI: −12.1%, −6.0%) for those identifying as Hispanic. Between 1999 and 2018, the uninsured prevalence gap between those identifying as White and those identifying as Black was reduced by 3.5% (95% CI: −5.8%, −1.1%), by 6.6% for those identifying as Asian (95% CI: −10.5%, −2.7%), and by 6.1% for those identifying as Hispanic (95% CI: −9.2%, −3.0%).

**Table 1.** Trends in Lack of Health Insurance by Race/Ethnicity and Study Periods.

| Race/Ethnicity | 1999-2009 |  | 2010-2015 |  | 2016-2018 |  | Delta 1999-2018 |
| --- | --- | --- | --- | --- | --- | --- | --- |
|  | Annualized Rate of Change, % (95% CI) | P Value | Annualized Rate of Change, % (95% CI) | P Value | Annualized Rate of Change, % (95% CI) | P Value | Absolute Difference, % (95% CI) |
| **White** |  |  |  |  |  |  |  |
| Prevalence | 0.3% (0.0%, 0.6%) | 0.05 | -0.9% (-1.3%, -0.5%) | <0.001 | -0.2% (-1.2%, 0.8%) | 0.72 | -2.9% (-3.6%, -2.2%) |
| **Black** |  |  |  |  |  |  |  |
| Prevalence | 0.4% (-0.3%, 1.1%) | 0.294 | -1.3% (-2.2%, -0.5%) | 0.002 | -0.6% (-4.7%, 3.5%) | 0.769 | -6.4% (-8.7%, -4.1%) |
| Gap w/ White | 0.1% (-0.3%, 0.4%) | 0.684 | -0.4% (-0.9%, 0%) | 0.052 | -0.5% (-3.0%, 2.0%) | 0.699 | -3.5% (-5.8%, -1.1%) |
| **Asian** |  |  |  |  |  |  |  |
| Prevalence | 0% (-0.8%, 0.8%) | 0.936 | -1.2% (-2.0%, -0.5%) | 0.002 | -0.7% (-3.2%, 1.8%) | 0.586 | -9.5% (-13.3%, -5.7%) |
| Gap w/ White | -0.4% (-0.7%, -0.1%) | 0.008 | -0.3% (-0.6%, 0.1%) | 0.136 | -0.7% (-2.3%, 0.9%) | 0.373 | -6.6% (-10.5%, -2.7%) |
| **Hispanic** |  |  |  |  |  |  |  |
| Prevalence | 0.6% (-0.2%, 1.3%) | 0.121 | -2.1% (-3.1%, -1.2%) | <0.001 | -1.2% (-5.3%, 2.9%) | 0.559 | -9.0% (-12.1%, -6.0%) |
| Gap w/ White | 0.2% (-0.2%, 0.6%) | 0.269 | -1.2% (-1.9%, -0.6%) | <0.001 | -1.1% (-4.5%, 2.2%) | 0.508 | -6.1% (-9.2%, -3.0%) |

**Figure 1.**
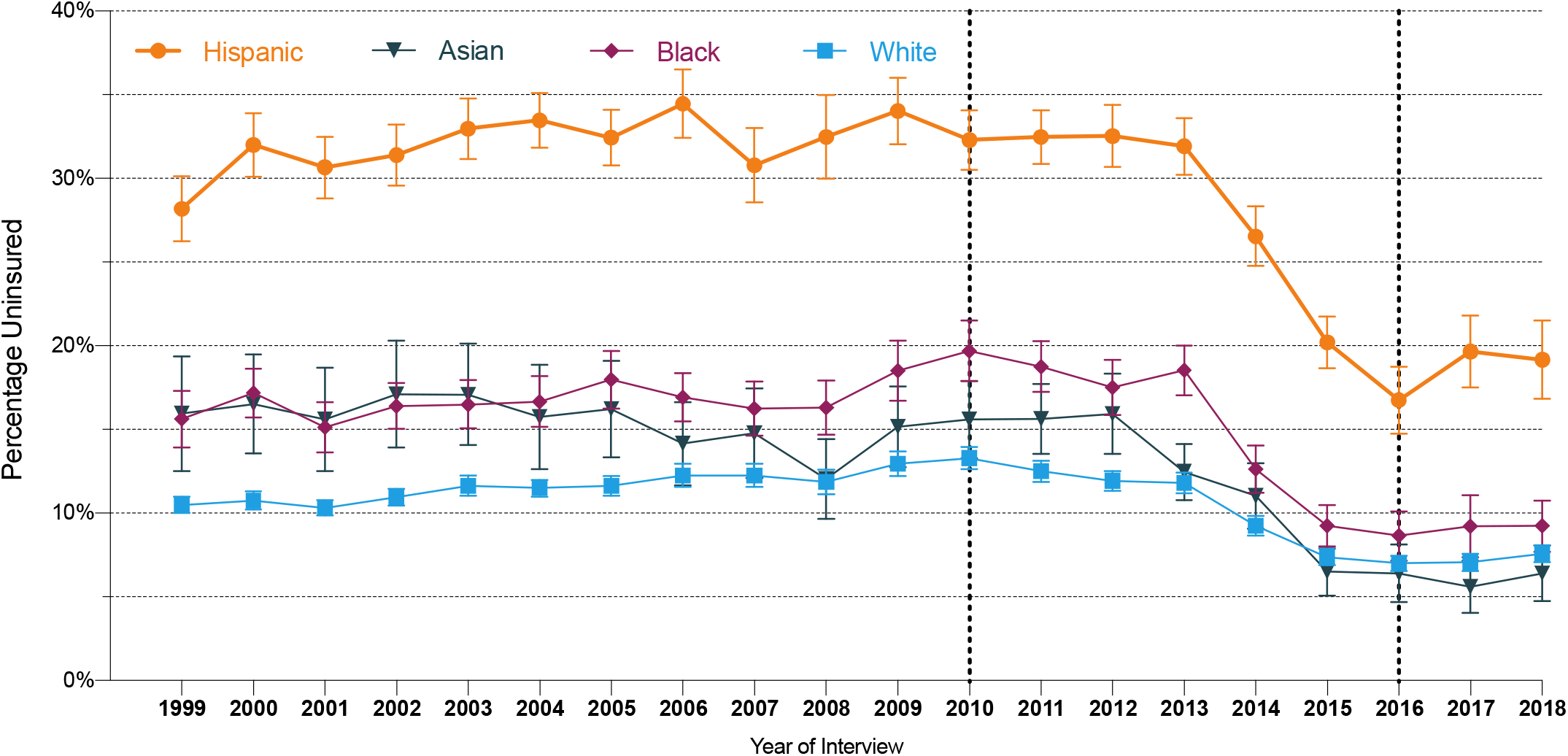
Annual age- and region-adjusted percentage of uninsured adults in the US by race/ethnicity, 1999 to 2018.

### Foregone or Delayed Medical Care Due to Cost

The age- and region-adjusted annual percentages of people who reported foregoing or delaying medical care due to cost by race/ethnicity are shown in **Figure 2**, and the annualized trends by study periods are shown in **Table 2**. The overall missing rate of foregone/delayed medical care due to cost status was 0.84%. From 1999 to 2009, overall rates increased for all 4 race/ethnicity groups. The slope was the steepest among those identifying as Hispanic (+1.1% per year, 95% CI: 0.75%, 1.4%), followed by those identifying as Black (+0.9% per year, 95% CI: 0.6%, 1.2%), those identifying as White (+0.7%, 95% CI: 0.5%, 0.8%), and those identifying as Asian (+0.3% per year, 95% CI: 0.01%, 0.6%). During this period, the gap between those identifying as White and those identifying as Hispanic and Black increased by 0.4% per year (95% CI: 0.3%, 0.5%) and 0.2% per year (95% CI: 0.1%, 0.4%), respectively. Between 2010 and 2015, a statistically significant decrease was observed for those identifying as White, Black, or Hispanic but not for those identifying as Asian. No significant trend was observed after 2016.

**Table 2.** Trends in Foregone or Delayed Medical Care Due to Cost by Race/Ethnicity and Study Periods.

| Race/Ethnicity | 1999-2009 |  | 2010-2015 |  | 2016-2018 |  | Delta 1999-2018<br>Absolute Difference,<br>% (95% CI) |
| --- | --- | --- | --- | --- | --- | --- | --- |
|  | Annualized Rate of<br>Change, % (95% CI) | P Value | Annualized Rate of<br>Change, % (95% CI) | P Value | Annualized Rate of<br>Change, % (95% CI) | P Value |  |
| **White** |  |  |  |  |  |  |  |
| Prevalence | 0.7% (0.5%, 0.9%) | <0.001 | -0.8% (-1.2%, -0.4%) | <0.001 | 0.4% (-0.4%, 1.3%) | 0.291 | 3.1% (2.3%, 3.9%) |
| **Black** |  |  |  |  |  |  |  |
| Prevalence | 0.9% (0.6%, 1.2%) | <0.001 | -0.8% (-1.3%, -0.3%) | 0.001 | -0.4% (-1.8%, 1.0%) | 0.55 | 3.1% (0.9%, 5.3%) |
| Gap w/ White | 0.2% (0.1%, 0.4%) | 0.002 | 0.0% (-0.3%, 0.2%) | 0.781 | -0.8% (-1.6%, -0.04%) | 0.04 | 0.1% (-2.3%, 2.4%) |
| **Asian** |  |  |  |  |  |  |  |
| Prevalence | 0.3% (0.01%, 0.6%) | 0.04 | -0.3% (-0.7%, -0.1%) | 0.199 | -0.4% (-1.4%, 0.6%) | 0.453 | 0.5% (-1.9%, 2.9%) |
| Gap w/ White | -0.4% (-0.5%, -0.2%) | <0.001 | 0.5% (0.3%, 0.7%) | <0.001 | -0.7% (-2.0%, 0.6%) | 0.283 | -2.5% (-5.1%, 0.0%) |
| **Hispanic** |  |  |  |  |  |  |  |
| Prevalence | 1.1% (0.8%, 1.4%) | <0.001 | -1.1% (-1.6%, -0.7%) | <0.001 | -0.5% (-1.7%, 0.7%) | 0.393 | 2.2% (0.4%, 4.0%) |
| Gap w/ White | 0.4% (0.3%, 0.5%) | <0.001 | -0.4% (-0.6%, -0.1%) | 0.005 | -0.7% (-1.2%, -0.2%) | 0.004 | -0.9% (-2.8%, 1.1%) |

**Figure 2.**
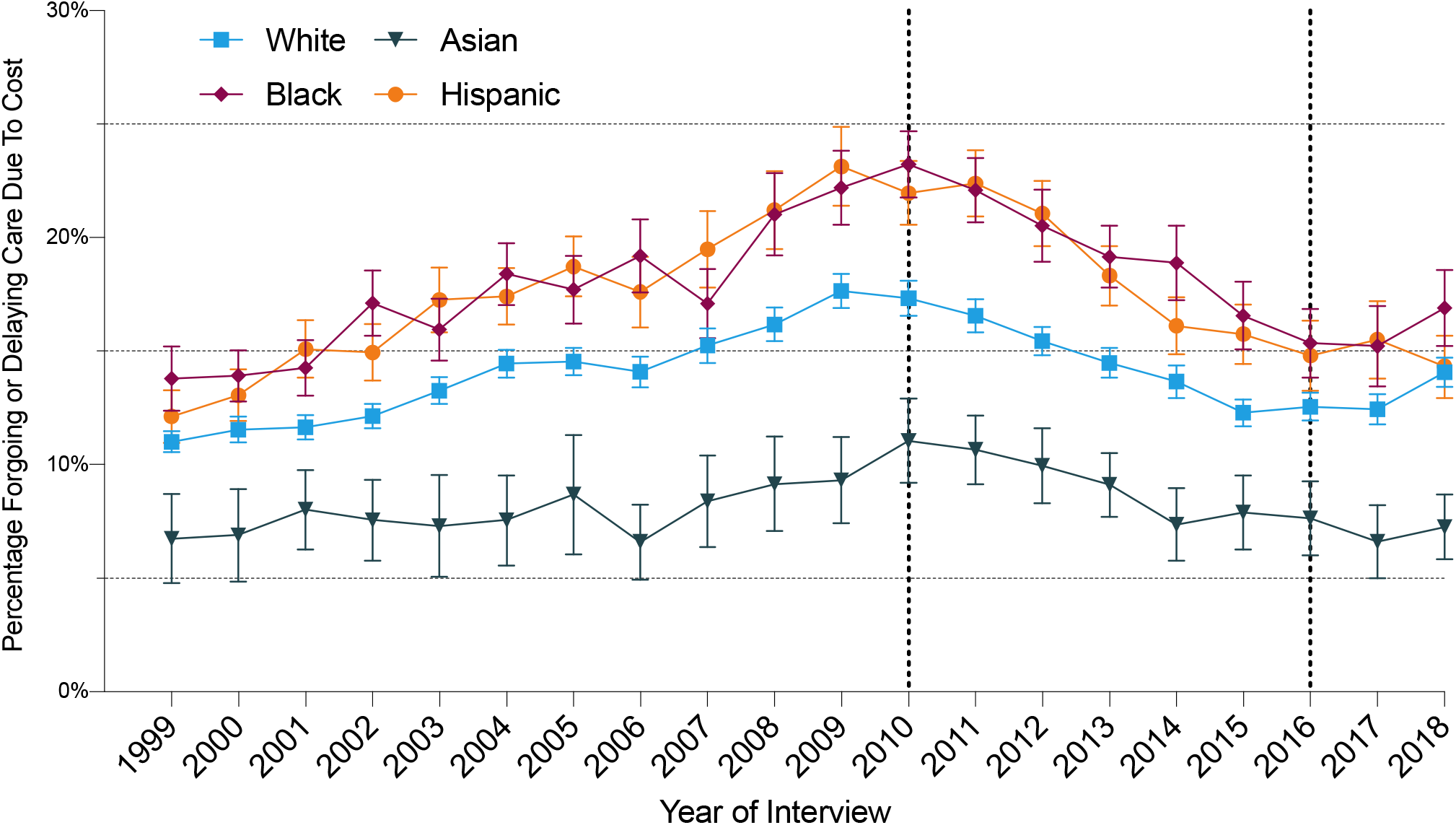
Annual age- and region-adjusted percentage of adults in the US who reported foregoing or delaying medical care due to cost in the past 12 months by race/ethnicity, 1999 to 2018.

Overall, between 1999 and 2018, the absolute difference in prevalence of foregone or delayed care due to cost was +3.1% for those identifying as White (95% CI: 2.3%, 3.9%), +3.1% for those identifying as Black (95% CI: 0.9%, 5.3%), +2.2% among those identifying as Hispanic (95% CI: 0.4%, 4.0%), and +0.5% among those identifying as Asian (95% CI: −2.0%, 2.9%), without any significant difference in disparities.

### No Usual Source of Care

The age- and region-adjusted annual percentages of people who did not have a usual source of care by race/ethnicity are shown in **Figure 3** and the annualized trends by study periods are shown in **Table 3**. The overall missing rate of having a usual source of care status was 0.76%. From 1999 to 2009, overall prevalence of this outcome increased slightly among those identifying as White (+0.2% per year, 95% CI: 0.1%, 0.3%) but remained stable for the other 3 subgroups. Between 2010 and 2015, the prevalence of this outcome decreased by 1.2% per year among those identifying as Hispanic (95% CI: −1.7%, −0.06%), and there were no significant trends among the other subgroups. No significant trend was identified in prevalence for any race/ethnicity subgroup between 2016–2018. Overall, between 1999 and 2018, the absolute change in prevalence of this outcome was −0.9% among those identifying as White (95% CI: −1.7%, 0.0%), −0.8% among those identifying as black (95% CI: −3.3%, 1.7%), −6.4% among those identifying as Asian (95% CI: −10.5%, −2.2%), and −4.9% among those identifying as Hispanic (95% CI: −7.6%, −2.2%). Similarly, during the study period, the gap between those identifying as White and Hispanic was reduced by 4 percentage points (95% CI: −6.8%, −1.2%).

**Table 3.** Trends in Lack of a Usual Source of Care by Race/Ethnicity and Study Periods.

| Race/Ethnicity | 1999-2018 |  | 2010-2015 |  | 2016-2018 |  | Delta 1999-2018 |
| --- | --- | --- | --- | --- | --- | --- | --- |
|  | Annualized Rate of Change, % (95% CI) | P Value | Annualized Rate of Change, % (95% CI) | P Value | Annualized Rate of Change, % (95% CI) | P Value | Absolute Difference, % (95% CI) |
| **White** |  |  |  |  |  |  |  |
| Prevalence | 0.2% (0.02%, 0.3%) | 0.007 | -0.1% (-0.5%, 0.2%) | 0.437 | 0.0% (-1.3%, 1.1%) | 0.905 | -0.9% (-1.7%, 0.0%) |
| **Black** |  |  |  |  |  |  |  |
| Prevalence | 0.3% (0.0%, 0.5%) | 0.083 | -0.4% (-1.2%, 0.5%) | 0.397 | 0.2% (-1.6%, 2.0%) | 0.848 | -0.8% (-3.3%, 1.7%) |
| Gap w/ White | 0.1% (0.0%, 0.2%) | 0.105 | -0.2% (-0.5%, 0.0%) | 0.157 | 0.1% (-0.6%, 0.8%) | 0.798 | 0.1% (-2.6%, 2.7%) |
| **Asian** |  |  |  |  |  |  |  |
| Prevalence | -0.1% (-0.4%, 0.1%) | 0.289 | -0.1% (-0.9%, 0.8%) | 0.85 | -0.8% (-3.1%, 1.5%) | 0.515 | -6.4% (-10.5%, -2.3%) |
| Gap w/ White | -0.3% (-0.6%, -0.02%) | 0.035 | 0.1% (-0.7%, 0.8%) | 0.892 | -0.7% (-2.9%, 1.5%) | 0.542 | -5.5% (-9.7%, -1.3%) |
| **Hispanic** |  |  |  |  |  |  |  |
| Prevalence | 0.3% (-0.2%, 0.7%) | 0.228 | -1.2% (-1.7%, -0.6%) | <0.001 | -0.4% (-2.5%, 1.7%) | 0.72 | -4.9% (-7.6%, -2.2%) |
| Gap w/ White | 0.1% (-0.1%, 0.4%) | 0.326 | -1.0% (-1.6%, -0.5%) | <0.001 | -0.3% (-2.9%, 1.5%) | 0.542 | -4.0% (-6.8%, -1.2%) |

**Figure 3.**
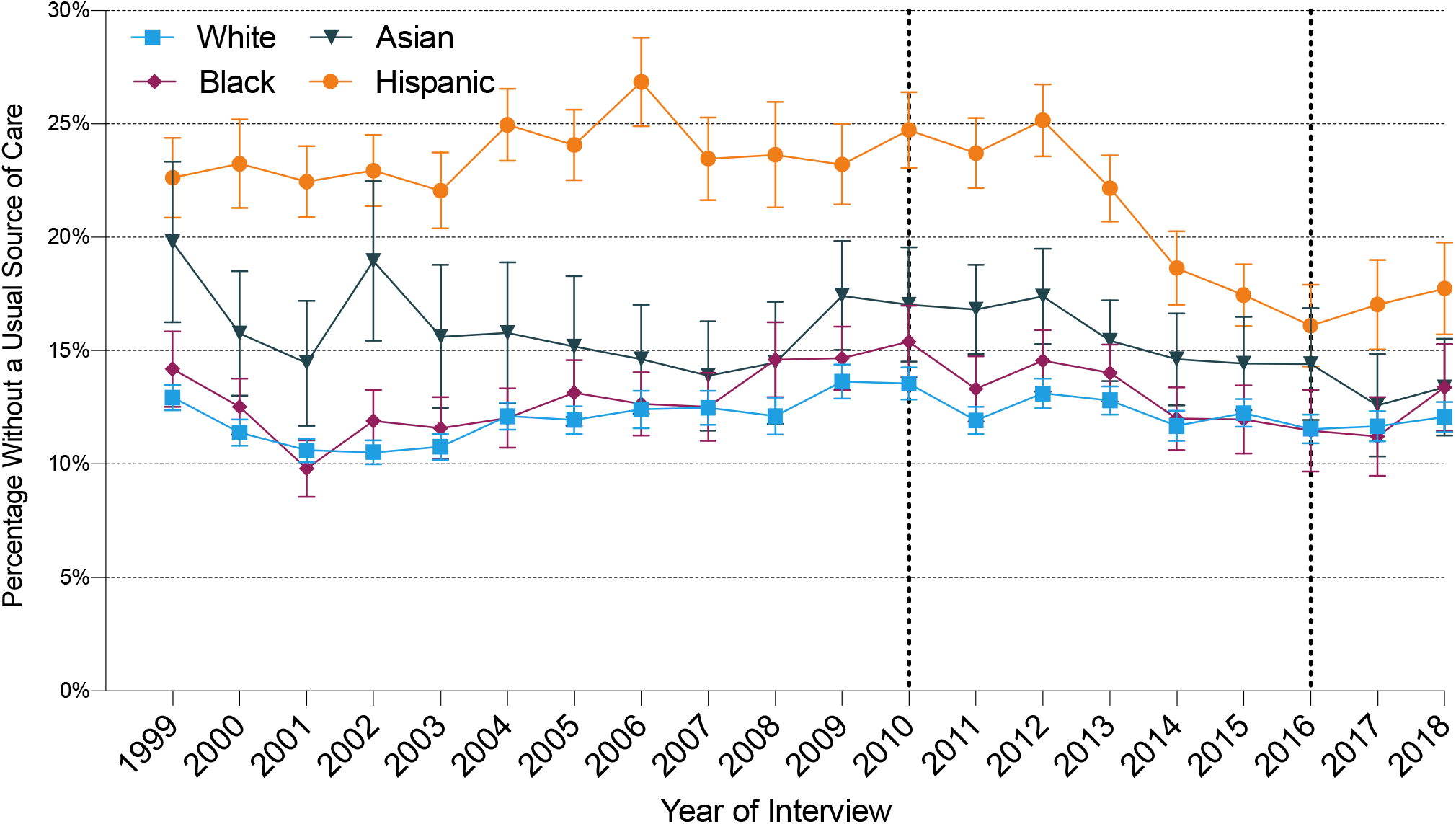
Age- and region-adjusted annual percentage of adults in the US without a usual source of care by race/ethnicity, 1999 to 2018.

## DISCUSSION

In this study, we report that over the past 20 years there have been modest improvements in insurance coverage, mostly during the last decade. However, in parallel there have been only slight reductions in the percentage of people without a usual source of care and no substantive reductions in the percentage of people forgoing or delaying care due to cost. Importantly, there have also been no substantial reductions in access to care disparities for Black and Hispanic individuals compared with White individuals. Particularly, the gap in foregone or delayed medical care has persisted despite the recent improvements in insurance coverage due to the ACA, indicating longstanding underinsurance. Of note, during this period total and per capita health care costs have nearly doubled.^27,28^

Our study expands the literature in two major ways. First, we report a comprehensive 20- year perspective of insurance coverage and access to care in the US by race and ethnicity. Several other studies have studied trends by race or ethnicity, but for shorter time frames that may limit the interpretation of change relative to historical levels. For example, Chen and colleagues^1^ found that in 2014, after the ACA was implemented, the rates of foregoing or delaying medical care due to cost was reduced for all races, narrowing the gaps between them. However, we found that, relative to 1999, rates of this outcome have actually increased, and disparities remained in 2018. The rate of foregoing care due to cost increased after 2008, coinciding with the economic recession,^20,22^ and decreased after 2011, but not to its 1999 level. This shows that even after the ACA was implemented, which reduced the rates of foregoing care due to cost in the states that implemented it,^29^ access to care has not substantially changed from what it was at the beginning of the century. Second, we found that after 2016, no significant gains were achieved in access to health care or its associated racial and ethnic disparities. Although data from only 3 years were available for analysis, which may limit our ability to draw conclusions on general trends during this period, we show that under the current US administration, access to health care services has plateaued or worsened for all race and ethnic groups. This could be explained by the continuous efforts to undermine the provisions of the ACA since 2017 which, although still limited, have already succeeded in decreasing Medicaid coverage in some states with increased barriers to access to care.^30-32^

Our study has important public health implications. The lack of improvement has occurred despite massive increases in health care spending. Also, the improvements in insurance coverage have not been accompanied by commensurate improvements in access to health care. Reducing the proportion of people who cannot get care when needed, and reducing racial/ethnic and income disparities, were objectives of Healthy People 2010 and 2020, and were included again in Healthy People 2030.^11,12,33^ Despite this, we found that no advancement was achieved in the past 2 decades among all 4 major race/ethnicity groups. Similarly, a series of Gallup surveys^34^ report that since 2001, there has been an increase in the proportion of families in the US with members who delay care due to cost, particularly among low-income families. They also report that in 2019, nearly 1 in 3 families in the US delayed any care due to cost and nearly 1 in 4 delayed care for a serious condition for the same reason.^34^ Together, these findings underscore the urgent need to increase not only insurance coverage but also the adequacy of the insurance that is being gained.

These findings do not imply that implementation of the ACA was not helpful. Previously uninsured individuals who enroll in Medicaid report lower out-of-pocket payments and lower prevalence of unmet medical needs due to cost, and are more likely to have a usual source of care and to have less psychological distress than those who remain uninsured.^35^ However, our findings also suggest that coverage alone may not be sufficient to completely protect individuals from experiencing barriers to access and to eliminate racial/ethnic disparities in access to health care. Despite increased coverage, little change was achieved in rates of unmet medical needs due to cost and lack of usual source of care. In fact, the proportion of inadequately insured adults (i.e., those with out-of-pocket expenditures >10% of their household income, or >5% if they are part of a low-income family) increased after the ACA,^36^ which could explain our findings. People with unmet medical needs due to cost or without a usual source of care experience higher health costs and worse health outcomes,^37-42^ and our findings indicate that these barriers are disproportionately more prevalent among racial and ethnic minorities.

In our analysis, Black and Hispanic respondents had the worst access to care and no substantive improvement over the 20-year timespan. It is not possible to fully address this issue without attending to the structural racism in our society. Although we used data up to 2018, the disproportionate burden of the COVID-19 pandemic on minority communities has likely worsened what we have observed.^43-46^

This study has several limitations. First, the data are self-reported. However, the outcomes and variables in our study reflect the actual experience of individuals with the health care system. Second, because of small numbers, we did not analyze race groups with smaller representation such as Alaskan Native, American Indian, or those not identifying with a single primary race. Third, although we used 3 of the major barriers to access to the health care system (insurance coverage, cost of care, and usual source of care), we did not include other structural barriers within the system such as language, transportation, or waiting times, which are also worth exploring. Fourth, although the association of the outcomes in our study and clinical outcomes has been described, we cannot directly make such an association.

In summary, 35 years after the Heckler Report, marked disparities in access to health care in the US persist, with individuals identifying as non-Hispanic Black and Hispanic continuing to experience more barriers to health care services compared with non-Hispanic White individuals. We failed to identify any major progress in reducing disparities and report substantial percentages of individuals in all groups who experience barriers to access to health care.

## Data Availability

We used publicly available data from the National Health Interview Survey using the Integrated Public Use Microdata Series (IPUMS) Health Surveys website.

https://nhis.ipums.org/nhis/

## ACKNOWLEDGEMENTS

### Funding

None.

### Disclosures

In the past three years, Dr. Krumholz received expenses and/or personal fees from UnitedHealth, IBM Watson Health, Element Science, Aetna, Facebook, the Siegfried and Jensen Law Firm, Arnold and Porter Law Firm, Martin/Baughman Law Firm, F-Prime, and the National Center for Cardiovascular Diseases in Beijing. He is an owner of Refactor Health and HugoHealth, and had grants and/or contracts from the Centers for Medicare & Medicaid Services, Medtronic, the U.S. Food and Drug Administration, Johnson & Johnson, and the Shenzhen Center for Health Information. Dr. Murugiah works under contract with the Centers for Medicare & Medicaid Services to support quality measurement programs. Dr. Lu is supported by the National Heart, Lung, and Blood Institute (K12HL138037) and the Yale Center for Implementation Science. She was a recipient of a research agreement, through Yale University, from the Shenzhen Center for Health Information for work to advance intelligent disease prevention and health promotion. Dr. Roy is a consultant for the Institute for Healthcare Improvement. The other co-authors report no potential competing interests.

## Notes

### Funding Statement

No external funding was received for this study.

### Author Declarations

This study received exemption for review from the Institutional Review Board at Yale University as NHIS data are publicly available and deidentified.

## REFERENCES

1. Chen J, Vargas-Bustamante A, Mortensen K, Ortega AN. Racial and ethnic disparities in health care access and utilization under the Affordable Care Act. Med Care. 2016;54(2):140–146.

2. Chou CH, Tulolo A, Raver EW, Hsu CH, Young G. Effect of race and health insurance on health disparities: results from the National Health Interview Survey 2010. J Health Care Poor Underserved. 2013;24(3):1353–1363.

3. Monheit AC, Vistnes JP. Race/ethnicity and health insurance status: 1987 and 1996. Med Care Res Rev. 2000;57 Suppl 1:11–35.

4. Manuel JI. Racial/ethnic and gender disparities in health care use and access. Health Serv Res. 2018;53(3):1407–1429.

5. 2018 National Healthcare Quality and Disparities Report. Rockville, MD: U.S. Department of Health and Human Services; 2019.

6. QuickStats: Percentage of adults aged 18-64 years with a usual place for health care, by race/ethnicity -National Health Interview Survey, United States, 2008 and 2018. MMWR Morb Mortal Wkly Rep. 2020;69(5):147.

7. Mayberry RM, Mili F, Ofili E. Racial and ethnic differences in access to medical care. Med Care Res Rev. 2000;57 Suppl 1:108–145.

8. Wang TF, Shi L, Nie X, Zhu J. Race/ethnicity, insurance, income and access to care: the influence of health status. Int J Equity Health. 2013;12:29.

9. United States Department of Health and Human Services Task Force on Black and Minority Health. Report of the Secretary’s Task Force on Black and Minority Health. 1985. [Accessed on July 30, 2020] Available from: http://resource.nlm.nih.gov/8602912.

10. Institute of Medicine Committee on Understanding and Eliminating Racial and Ethnic Disparities in Health Care. Unequal Treatment: Confronting Racial and Ethnic Disparities in Health Care. National Academies Press (US) Copyright 2002 by the National Academy of Sciences. Washington (DC). 2003.

11. U.S. Department of Health and Human Services. Healthy People 2010. November 2000. [Accessed on October 20, 2020] Available from: https://www.healthypeople.gov/2010/Document/html/uih/uih_2.htm#goals.

12. U.S. Department of Health and Human Services, Office of Disease Prevention and Health Promotion. Healthy People 2020. 2010. [Accessed on October 20, 2020] Available from: https://www.healthypeople.gov/2020/topics-objectives/topic/Access-to-Health-Services.

13. Koh HK, Graham G, Glied SA. Reducing racial and ethnic disparities: the action plan from the Department of Health and Human Services. Health Aff (Millwood). 2011;30(10):1822–1829.

14. Centers for Disease Control and Prevention. About the National Health Interview Survey. 2019. [Accessed on October 6, 2020] Available from: https://www.cdc.gov/nchs/nhis/about_nhis.htm.

15. Blewett LA, Rivera Drew JA, King ML and Williams KCW. IPUMS Health Surveys: National Health Interview Survey, Version 6.4. Minneapolis, MN: IPUMS, 2019. https://doi.org/10.18128/D070.V6.4.

16. IRB Exemption. [Accessed on October 23, 2020] Available from: https://www.hhs.gov/ohrp/regulations-and-policy/decision-charts-2018/index.html.

17. National Center for Health Statistics. Evaluation and Editing of Health Insurance Data, National Health Insurance Survey. June 14, 2018. [Accessed on October 10, 2020] Available from: https://www.cdc.gov/nchs/nhis/health_insurance/hi_eval.htm.

18. National Center for Health Statistics. Data Brief: Health care access and utilization among adults aged 18-64, by race and Hispanic origin: United States, 2013 and 2014. 2015. [Accessed on July 1, 2020] Available from: https://www.cdc.gov/nchs/products/databriefs/db208.htm.

19. Caraballo C, Valero-Elizondo J, Khera R, et al. Burden and consequences of financial hardship from medical bills among nonelderly adults with diabetes mellitus in the United States. Circ Cardiovasc Qual Outcomes. 2020;13(2):e006139.

20. Travers JL, Cohen CC, Dick AW, Stone PW. The Great American Recession and forgone healthcare: Do widened disparities between African-Americans and Whites remain? PLoS One. 2017;12(12):e0189676.

21. Johnson PJ, Jou J, Upchurch DM. Health care disparities among U.S. women of reproductive age by level of psychological distress. J Womens Health (Larchmt). 2019;28(9):1286–1294.

22. Burgard SA, Hawkins JM. Race/Ethnicity, educational attainment, and foregone health care in the United States in the 2007-2009 recession. Am J Public Health. 2014;104(2):e134–140.

23. Mszar R, Grandhi GR, Valero-Elizondo J, et al. Cumulative burden of financial hardship from medical bills across the spectrum of diabetes mellitus and atherosclerotic cardiovascular disease among non-elderly adults in the United States. J Am Heart Assoc. 2020;9(10):e015523.

24. United States Census Bureau. People in families by family structure, age, and sex, iterated by income-to-poverty ratio and race. 2018. Available from: https://www.census.gov/

25. Khera R, Valero-Elizondo J, Das SR, et al. Cost-related medication nonadherence in adults with atherosclerotic cardiovascular disease in the United States, 2013 to 2017. Circulation. 2019;140(25):2067–2075.

26. NHIS data, questionnaires and related documentation. 2019. [Accessed on October 11, 2020] Available from: https://www.cdc.gov/nchs/nhis/data-questionnaires-documentation.htm.

27. Centers for Medicare & Medicaid Services. National Health Expenditure Data, Historical. December 2019. [Accessed on October 6, 2020] Available from: https://www.cms.gov/Research-Statistics-Data-and-Systems/Statistics-Trends-and-Reports/NationalHealthExpendData/NationalHealthAccountsHistorical.

28. Kamal R, McDermott D, Cox C. Peterson Center on Healthcare-Kaiser Family Foundation Health System Tracker. How has U.S. spending on healthcare changed over time? December 2019. [Accessed on October 6, 2020] Available from: https://www.healthsystemtracker.org/chart-collection/u-s-spending-healthcare-changed-time/.

29. Miller S, Wherry LR. Health and access to care during the first 2 years of the ACA Medicaid expansions. N Engl J Med. 2017;376(10):947–956.

30. Chen L, Sommers BD. Work requirements and Medicaid disenrollment in Arkansas, Kentucky, Louisiana, and Texas, 2018. Am J Public Health. 2020;110(8):1208–1210.

31. Sommers BD, Chen L, Blendon RJ, Orav EJ, Epstein AM. Medicaid work requirements in Arkansas: two-year impacts on coverage, employment, and affordability of care. Health Aff (Millwood). 2020;39(9):1522–1530.

32. Sommers BD, Goldman AL, Blendon RJ, Orav EJ, Epstein AM. Medicaid work requirements-results from the first year in Arkansas. N Engl J Med. 2019;381(11):1073–1082.

33. U.S. Department of Health and Human Services, Office of Disease Prevention and Health Promotion. Healthy People 2030. [Accessed on October 5, 2020] Available from: https://health.gov/healthypeople/objectives-and-data/browse-objectives/health-care-access-and-quality/reduce-proportion-people-who-cant-get-medical-care-when-they-need-it-ahs-04/data.

34. Saad L. More Americans delaying medical treatment due to cost. Gallup. December 9, 2019. [Accessed on October 3, 2020] Available from: https://news.gallup.com/poll/269138/americans-delaying-medical-treatment-due-cost.aspx.

35. Winkelman TNA, Segel JE, Davis MM. Medicaid enrollment among previously uninsured Americans and associated outcomes by race/ethnicity-United States, 2008-2014. Health Serv Res. 2019;54 Suppl 1(Suppl 1):297–306.

36. Collins SR, Bhupal HK, Doty MM. Health insurance coverage eight years after the ACA: fewer uninsured Americans and shorter coverage gaps, but more underinsured. Commonwealth Fund. 2019;7. [Accessed on October 28, 2020] Available from: https://collections.nlm.nih.gov/catalog/nlm:nlmuid-101750766-pdf.

37. De Maeseneer JM, De Prins L, Gosset C, Heyerick J. Provider continuity in family medicine: does it make a difference for total health care costs? Ann Fam Med. 2003;1(3):144–148.

38. Ettner SL. The timing of preventive services for women and children: the effect of having a usual source of care. Am J Public Health. 1996;86(12):1748–1754.

39. Erskine NA, Gandek B, Tran HV, et al. Barriers to healthcare access and to improvements in health-related quality of life after an acute coronary syndrome (TRACE-CORE). Am J Cardiol. 2018;122(7):1121–1127.

40. Erskine NA, Waring ME, McManus DD, Lessard D, Kiefe CI, Goldberg RJ. Barriers to healthcare access and long-term survival after an acute coronary syndrome. J Gen Intern Med. 2018;33(9):1543–1550.

41. Fullerton CA, Witt WP, Chow CM, et al. Impact of a usual source of care on health care use, spending, and quality among adults with mental health conditions. Adm Policy Ment Health. 2018;45(3):462–471.

42. Lee W, Lloyd JT, Giuriceo K, Day T, Shrank W, Rajkumar R. Systematic review and meta-analysis of patient race/ethnicity, socioeconomics, and quality for adult type 2 diabetes. Health Serv Res. 2020.

43. National Center for Health Statistics. Health disparities: race and Hispanic origin, provisional death counts for coronavirus disease 2019 (COVID-19). 2020. Accessed on August 12, 2020. Avaliable from: https://www.cdc.gov/nchs/nvss/vsrr/covid19/health_disparities.htm.

44. Rossen LM, Branum AM, Ahmad FB, Sutton P, Anderson RN. Excess deaths associated with COVID-19, by age and race and ethnicity—United States, January 26–October 30 2020. Centers for Disease Control and Prevention, Morb Mortal Wkly Rep. October 20, 2020. [Accessed on October 20, 2020] Available from: https://www.cdc.gov/mmwr/volumes/69/wr/pdfs/mm6942e2-H.pdf.

45. Banthin J, Blumberg LJ, Simpson M, Buettgens M, Wang R. Changes in health insurance coverage due to the COVID-19 recession: preliminary estimates using microsimulation. Washington, DC: Urban Institute. 2020.

46. The National Center For Coverage Innovation. The COVID-19 pandemic and resulting economic crash have caused the greatest health insurance losses in American history. July 17, 2020. Accessed on August 13, 2020. Available from: https://www.familiesusa.org/wp-content/uploads/2020/07/COV-254_Coverage-Loss_Report_7-17-20.pdf.

