## Supplemental Material for "Racial and Ethnic Disparities in Access to Health Care Among Adults in the United States: A 20-Year National Health Interview Survey Analysis, 1999–2018"

**SUPPLEMENTARY MATERIAL**

**Online Table S1**. General Characteristics of Study Population by Race/Ethnicity in 1999–2000 and 2017–2018.

**Online Figure S1**. Study Population Flowchart.

**Online Figure S2**. Race and Ethnicity Distribution Among Adults in the US, National Health Interview Survey 1999-2018.

**Table S1. General Characteristics of Study Population by Race/Ethnicity in 1999–2000 and 2017–2018.**

| **Characteristics** | **Non-Hispanic White** | | **Hispanic** | | **Non-Hispanic Black** | | **Non-Hispanic Asian** | |
| --- | --- | --- | --- | --- | --- | --- | --- | --- |
|  | **1999–2000** | **2017–2018** | **1999–2000** | **2017–2018** | **1999–2000** | **2017–2018** | **1999–2000** | **2017–2018** |
|  | **n (%, 95% CI)** | **n (%, 95% CI)** | **n (%, 95% CI)** | **n (%, 95% CI)** | **n (%, 95% CI)** | **n (%, 95% CI)** | **n (%, 95% CI)** | **n (%, 95% CI)** |
| **Total** | 41,882 | 36,410 | 10,305 | 6,423 | 8,758 | 5,884 | 1,597 | 2,640 |
| **Age (years)** |  |  |  |  |  |  |  |  |
| 18**–**39 | 15,169 (39.5% [38.9–40.2]) | 10,050 (33.3% [32.6–34.1]) | 3,610 (37.1% [35.0–39.2]) | 2,886 (50.1% [48.4–51.8]) | 3,862 (49.8% [48.3–51.3]) | 1,994 (43.7% [41.8–45.6]) | 844 (52.4% [49.2–55.6]) | 1,085 (41.9% [39.3–44.6]) |
| 40**–**64 | 17,246 (42.2% [41.5–42.8]) | 14,919 (42.3% [41.6–43.0]) | 6,617 (62.1% [60.0–64.2]) | 2,492 (39.1% [37.6–40.6]) | 3,539 (38.5% [37.1–39.8]) | 2,434 (41.1% [39.3–42.9]) | 581 (38.3% [35.1–41.4]) | 1,005 (42.5% [40.2–44.9]) |
| ≥65 | 9,467 (18.3% [17.8–18.8]) | 11,441 (24.4% [23.7–25.1]) | 78 (0.8% [0.6–1.1]) | 1,045 (10.8% [9.9–11.8]) | 1,357 (11.8% [11.0–12.6]) | 1,456 (15.3% [14.2–16.3]) | 172 (9.4% [7.8–11.4]) | 550 (15.5% [13.9–17.4]) |
| **Sex** |  |  |  |  |  |  |  |  |
| Men | 18,512 (48.1% [47.6–48.7]) | 16,798 (48.6% [48.0–49.3]) | 4,399 (49.4% [48.2–50.6]) | 2,788 (49.7% [48.1–51.4]) | 3,255 (44.4% [43.0–45.9]) | 2,435 (45.5% [43.7–47.2]) | 726 (48.7% [45.4–51.9]) | 1,270 (46.7% [44.4–49.0]) |
| Women | 23,370 (51.9% [51.3–52.4]) | 19,612 (51.4% [50.7–52.0]) | 5,906 (50.6% [49.4–51.8]) | 3,635 (50.3% [48.7–51.9]) | 5,503 (55.6% [54.1–57.0]) | 3,449 (54.5% [52.8–56.3]) | 871 (51.4% [48.1–54.6]) | 1,370 (53.3% [51.1–55.6]) |
| **Citizenship status** |  |  |  |  |  |  |  |  |
| US citizen | 41,177 (98.3% [98.2–98.5]) | 35,943 (98.4% [98.2–98.6]) | 6,617 (62.1% [60.0–64.2]) | 4,694 (71.5% [69.8–73.3]) | 8,385 (95.2% [94.5–95.8]) | 5,655 (94.8% [93.6–95.8]) | 921 (57.1% [53.5–60.6]) | 1,844 (69.9% [67.2–72.4]) |
| Non-US citizen | 672 (1.6% [1.5–1.8]) | 446 (1.5% [1.3–1.7]) | 3,610 (37.1% [35.0–39.2]) | 1,664 (27.4% [25.7–29.1]) | 360 (4.6% [4.0–5.3]) | 220 (5.1% [4.1–6.2]) | 658 (41.6% [38.0–45.2]) | 775 (29.2% [26.7–31.8]) |
| Unknown | 33 (0.0% [0.0–0.0]) | 21 (0.0% [0.0–0.1]) | 78 (0.8% [0.6–1.1]) | 65 (1.1% [0.8–1.6]) | 13 (0.2% [0.0–0.4]) | 9 (0.1% [0.0–0.3]) | 18 (1.3% [0.7–2.4]) | 21 (0.9% [0.5–1.7]) |
| **Education level** |  |  |  |  |  |  |  |  |
| Less than high school | 5,770 (13.3% [12.8–13.8]) | 2,708 (7.4% [6.9–7.9]) | 4,810 (44.0% [42.4–45.7]) | 1,831 (27.5% [25.7–29.4]) | 2,228 (24.1% [22.8–25.5]) | 948 (14.1% [12.8–15.6]) | 179 (11.2% [9.3–13.5]) | 221 (7.9% [6.6–9.4]) |
| High school diploma /GED | 12,918 (31.5% [30.8–32.1]) | 8,710 (23.6% [22.8–24.3]) | 2,394 (24.0% [22.9–25.0]) | 1,615 (26.4% [24.8–28.0]) | 2,569 (31.1% [29.8–32.4]) | 1,646 (28.7% [27.1–30.3]) | 276 (17.7% [15.8–19.8]) | 383 (15.2% [13.4–17.3]) |
| Some college | 12,147 (29.3% [28.8–29.8]) | 11,509 (31.3% [30.5–32.0]) | 2,086 (21.4% [20.2–22.6]) | 1,737 (27.8% [26.3–29.4]) | 2,558 (29.3% [28.0–30.6]) | 1,905 (32.7% [31.0–34.5]) | 367 (23.8% [21.3–26.5]) | 546 (20.6% [18.7–22.8]) |
| ≥Bachelor’s degree | 10,685 (25.2% [24.5–25.9]) | 13,386 (37.5% [36.4–38.6]) | 888 (9.2% [8.4–10.1]) | 1,193 (17.3% [15.9–18.8]) | 1,300 (14.3% [13.2–15.5]) | 1,347 (23.9% [22.0–25.9]) | 750 (45.6% [42.6–48.6]) | 1,478 (55.6% [52.5–58.6]) |
| Unknown | 362 (0.8% [0.7–0.9]) | 97 (0.3% [0.2–0.4]) | 127 (1.4% [1.1–1.8]) | 47 (1.0% [0.7–1.5]) | 103 (1.3% [1.0–1.6]) | 38 (0.6% [0.4–0.9]) | 25 (1.8% [1.2–2.7]) | 12 (0.7% [0.3–1.5]) |
| **Characteristics** | **Non-Hispanic White** | | **Hispanic** | | **Non-Hispanic Black** | | **Non-Hispanic Asian** | |
|  | **1999–2000** | **2017–2018** | **1999–2000** | **2017–2018** | **1999–2000** | **2017–2018** | **1999–2000** | **2017–2018** |
|  | **n (%, 95% CI)** | **n (%, 95% CI)** | **n (%, 95% CI)** | **n (%, 95% CI)** | **n (%, 95% CI)** | **n (%, 95% CI)** | **n (%, 95% CI)** | **n (%, 95% CI)** |
| **Family income** |  |  |  |  |  |  |  |  |
| High/middle income | 30,643 (77.0% [76.4–77.6]) | 27,211 (78.8% [77.9–79.6]) | 4,412 (49.3% [47.6–50.9]) | 3,165 (53.5% [51.4–55.5]) | 4,372 (54.5% [52.5–56.5]) | 2,961 (55.3% [52.9–57.6]) | 1,082 (71.8% [68.3–75.1]) | 1,886 (73.8% [71.0–76.5]) |
| Low income | 11,239 (23.0% [22.4–23.6]) | 9,199 (21.2% [20.4–22.1]) | 5,893 (50.7% [49.1–52.4]) | 3,258 (46.5% [44.5–48.6]) | 4,386 (45.5% [43.6–47.5]) | 2,923 (44.7% [42.4–47.1]) | 515 (28.2% [24.9–31.7]) | 754 (26.2% [23.5–29.0]) |
| **Insurance status** |  |  |  |  |  |  |  |  |
| Insured | 37,216 (88.7% [88.3–89.1]) | 34,050 (93.2% [92.8–93.6]) | 6,669 (63.4% [61.6–65.2]) | 4,937 (75.4% [73.4–77.2]) | 7,036 (78.9% [77.8–80.0]) | 5,216 (87.0% [85.6–88.3]) | 1,329 (82.3% [80.0–84.5]) | 2,469 (93.2% [91.8–94.3]) |
| Uninsured | 4,522 (11.0% [10.6–11.4]) | 2,270 (6.5% [6.2–6.9]) | 3,591 (36.1% [34.3–37.9]) | 1,454 (23.8% [22.0–25.8]) | 1,663 (20.1% [19.1–21.2]) | 632 (11.9% [10.7–13.4]) | 260 (17.3% [15.1–19.7]) | 163 (6.3% [5.2–7.6]) |
| Unknown | 144 (0.3% [0.3–0.4]) | 90 (0.3% [0.2–0.4]) | 45 (0.5% [0.4–0.7]) | 32 (0.8% [0.5–1.3]) | 59 (1.0% [0.7–1.3]) | 36 (1.1% [0.7–1.7]) | 8 (0.4% [0.2–0.9]) | 8 (0.6% [0.3–1.2]) |
| **Region** |  |  |  |  |  |  |  |  |
| Northeast | 8,168 (20.2% [19.5–21.0]) | 6,431 (19.2% [17.5–21.1]) | 1,639 (15.7% [14.5–17.1]) | 789 (13.5% [11.2–16.2]) | 1,582 (18.0% [16.7–19.4]) | 733 (16.0% [13.5–18.8]) | 333 (21.2% [18.7–24.0]) | 493 (20.8% [16.9–25.4]) |
| Midwest | 12,012 (29.4% [28.5–30.3]) | 10,273 (27.4% [25.7–29.3]) | 637 (7.9% [6.9–9.1]) | 640 (9.4% [7.6–11.6]) | 1,698 (18.8% [17.2–20.5]) | 914 (15.2% [13.0–17.8]) | 214 (14.7% [11.9–18.0]) | 325 (11.8% [9.5–14.6]) |
| South | 13,935 (33.9% [33.0–34.8]) | 11,884 (33.0% [30.9–35.2]) | 3,729 (35.4% [33.2–37.8]) | 2,614 (37.1% [32.5–42.0]) | 4,645 (55.7% [53.4–57.9]) | 3,754 (60.5% [56.7–64.3]) | 298 (18.9% [16.4–21.8]) | 683 (25.5% [21.6–30.0]) |
| West | 7,767 (16.5% [15.8–17.2]) | 7,822 (20.4% [18.3–22.7]) | 4,300 (40.9% [38.6–43.3]) | 2,380 (40.0% [35.3–44.8]) | 833 (7.6% [6.7–8.5]) | 483 (8.3% [6.9–9.9]) | 752 (45.2% [41.2–49.2]) | 1,139 (41.8% [36.7–47.1]) |
| **Marital status** |  |  |  |  |  |  |  |  |
| Unmarried | 20,259 (38.3% [37.5–39.0]) | 19,152 (43.6% [42.8–44.3]) | 4,953 (41.3% [40.2–42.5]) | 3,569 (50.6% [48.9–52.2]) | 6,232 (62.2% [60.8–63.7]) | 4,364 (67.2% [65.5–68.9]) | 664 (35.5% [32.5–38.6]) | 1,123 (34.9% [32.5–37.4]) |
| Married/Living with partner | 21,431 (61.4% [60.6–62.1]) | 17,179 (56.3% [55.5–57.0]) | 5,302 (58.3% [57.1–59.4]) | 2,840 (49.1% [47.5–50.7]) | 2,459 (37.1% [35.7–38.6]) | 1,502 (32.6% [30.9–34.3]) | 923 (64.0% [61.0–67.0]) | 1,509 (64.9% [62.4–67.4]) |
| Unknown | 192 (0.3% [0.3–0.4]) | 79 (0.2% [0.1–0.2]) | 50 (0.4% [0.3–0.5]) | 14 (0.4% [0.2–0.6]) | 67 (0.6% [0.5–0.9]) | 18 (0.3 [0.2–0.4]) | 10 (0.5% [0.3–0.9]) | 8 (0.2% [0.0–0.5]) |
| **Employment Status** |  |  |  |  |  |  |  |  |
| With a job/Working | 26,251 (65.6% [65.0–66.3]) | 20,990 (62.1% [61.3–62.9]) | 6,430 (66.1% [64.9–67.4]) | 4,075 (66.5% [64.7–68.3]) | 5,410 (63.9% [62.4–65.4]) | 3,248 (61.0% [59.0–63.1]) | 1,029 (65.9% [63.2–68.5]) | 1,667 (67.1% [64.9–69.3]) |
| **Characteristics** | **Non-Hispanic White** | | **Hispanic** | | **Non-Hispanic Black** | | **Non-Hispanic Asian** | |
|  | **1999-2000** | **2017-2018** | **1999-2000** | **2017-2018** | **1999-2000** | **2017-2018** | **1999-2000** | **2017-2018** |
|  | **n (%, 95% CI)** | **n (%, 95% CI)** | **n (%, 95% CI)** | **n (%, 95% CI)** | **n (%, 95% CI)** | **n (%, 95% CI)** | **n (%, 95% CI)** | **n (%, 95% CI)** |
| Not in labor force | 15,037 (32.9% [32.2–33.6]) | 14,581 (35.4% [34.6–36.2]) | 3,579 (31.1% [29.9–32.3]) | 2,138 (29.8% [28.0–31.8]) | 3,020 (31.9% [30.5–33.4]) | 2,308 (32.4% [30.5–34.3]) | 530 (31.7% [29.1–34.4]) | 907 (30.0% [27.8–32.3]) |
| Unemployed | 562 (1.4% [1.2–1.5]) | 824 (2.5% [2.3–2.7]) | 276 (2.5% [2.2–2.9]) | 208 (3.6% [3.0–4.3]) | 308 (3.9% [3.4–4.6]) | 324 (6.5% [5.7–7.4]) | 35 (2.2% [1.5–3.1]) | 64 (2.8% [2.1–3.7]) |
| Unknown | 32 (0.1% [0.1–0.1]) | 15 (0.0% [0.0–0.1]) | 20 (0.3% [0.2–0.4]) | 2 (0.02% [0.0–0.1]) | 20 (0.3% [0.2–0.4]) | 4 (0.1% [0.0–0.3]) | 3 (0.2% [0.0–0.8]) | 2 (0.1% [0.0–0.4]) |
| **Comorbidities** |  |  |  |  |  |  |  |  |
| Hypertension | 10,466 (23.0% [22.5–23.4]) | 13,280 (32.7% [31.9–33.4]) | 1,762 (15.3% [14.4–16.2]) | 1,673 (22.3% [21.0–23.8]) | 2,857 (28.8% [27.6–30.1]) | 2,687 (37.6 [35.9–39.3]) | 237 (14.9% [12.9–17.2]) | 670 (24.2% [22.2–26.3]) |
| Diabetes | 2,303 (5.2% [5.0–5.4]) | 3,698 (9.1% [8.7–9.4]) | 720 (6.3% [5.8–6.9]) | 766 (10.5% [9.6–11.4]) | 803 (8.2% [7.5–8.9]) | 888 (11.7% [10.8–12.7]) | 57 (3.9% [3.0–5.1]) | 247 (8.9% [7.7–10.3]) |
| Prior stroke/MI | 2,404 (5.1% [4.9–5.3]) | 2,762 (6.4% [6.1–6.8]) | 307 (2.5% [2.2–2.9]) | 266 (3.3% [2.8–3.9]) | 456 (4.5% [4.0–5.1]) | 484 (6.0% [5.4–6.7]) | 35 (1.7% [1.2–2.2]) | 97 (3.2% [2.5–4.0]) |
| Cancer | 3,628 (7.8% [7.5–8.1]) | 5,164 (12.3 [11.9–12.8]) | 235 (2.2% [1.9–2.5]) | 289 (3.4% [2.9–3.8]) | 301 (2.9% [2.5–3.4]) | 379 (4.8% [4.3–5.4]) | 31 (1.7% [1.1–2.7]) | 126 (4.3% [3.4–5.3]) |
| Emphysema / Chronic bronchitis | 2,785 (6.2% [5.9–6.5]) | 2,163 (5.3% [5.0–5.6]) | 330 (2.9% [2.5–3.4]) | 231 (2.8% [2.4–3.3]) | 444 (4.3% [3.8–4.7]) | 332 (4.4% [3.7–5.1]) | 35 (1.6% [1.1–2.4]) | 58 (2.0% [1.4–2.7]) |
| **Current smoker** | 10,085 (24.2% [23.7–24.8]) | 5,712 (15.1% [14.6–15.7]) | 1,915 (18.3% [17.4–19.3]) | 690 (9.8% [9.0–10.8]) | 2,124 (23.8% [22.6–25.1]) | 973 (14.7% [13.5–16.1]) | 247 (14.9% [12.9–17.2]) | 201 (7.2% [6.0–8.5]) |
| **Flu vaccine in past 12m** | 13,642 (30.7% [30.0–31.3]) | 17,708 (47.1% [46.4–47.9]) | 1,982 (18.6% [17.5–19.6]) | 2,379 (34.9% [33.3–36.5]) | 1,879 (20.6% [19.7–21.5]) | 2,198 (35.2% [33.4–37.1]) | 426 (26.1% [23.3–29.2]) | 1,270 (48.2% [45.6–50.7]) |
| **Obese (BMI ≥30 kg/m^2^)** | 8,006 (20.2% [19.7–20.6]) | 10,627 (30.3% [29.6–31.0]) | 2,339 (23.0% [21.9–24.1]) | 2,093 (34.0% [32.5–35.6]) | 2,542 (29.1% [28.0–30.2]) | 2,312 (39.4% [37.8–41.1]) | 84 (6.1% [4.9–7.7]) | 289 (12.0% [10.5–13.6]) |
| Abbreviations: BMI, body mass index; CI, confidence interval; MI, myocardial infarction; US, United States | | | | | | | | |

**Figure S1. Study Population Flowchart.**
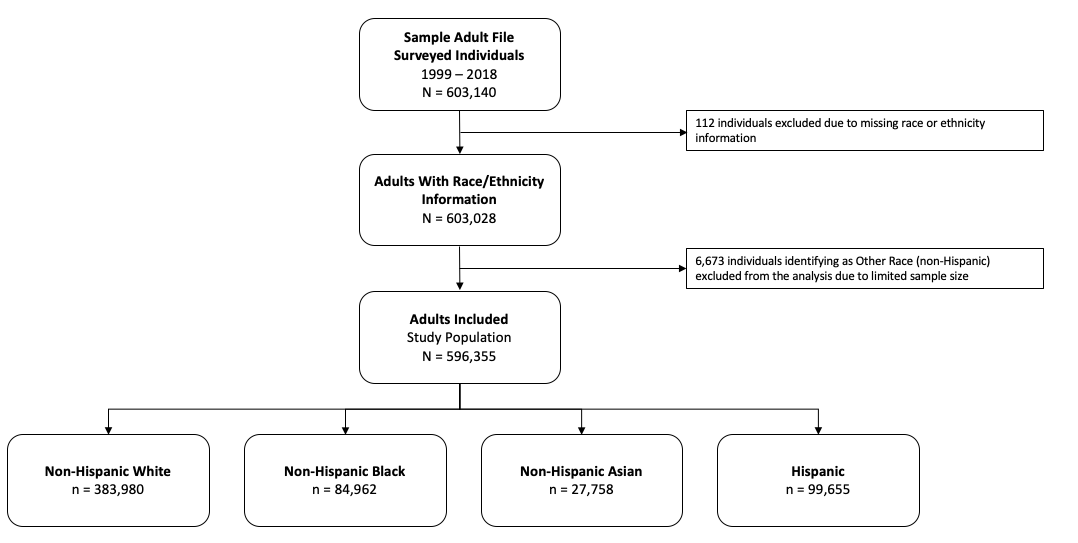

**Figure S2. Race and Ethnicity Distribution Among Adults in the US, National Health Interview Survey 1999-2018.**

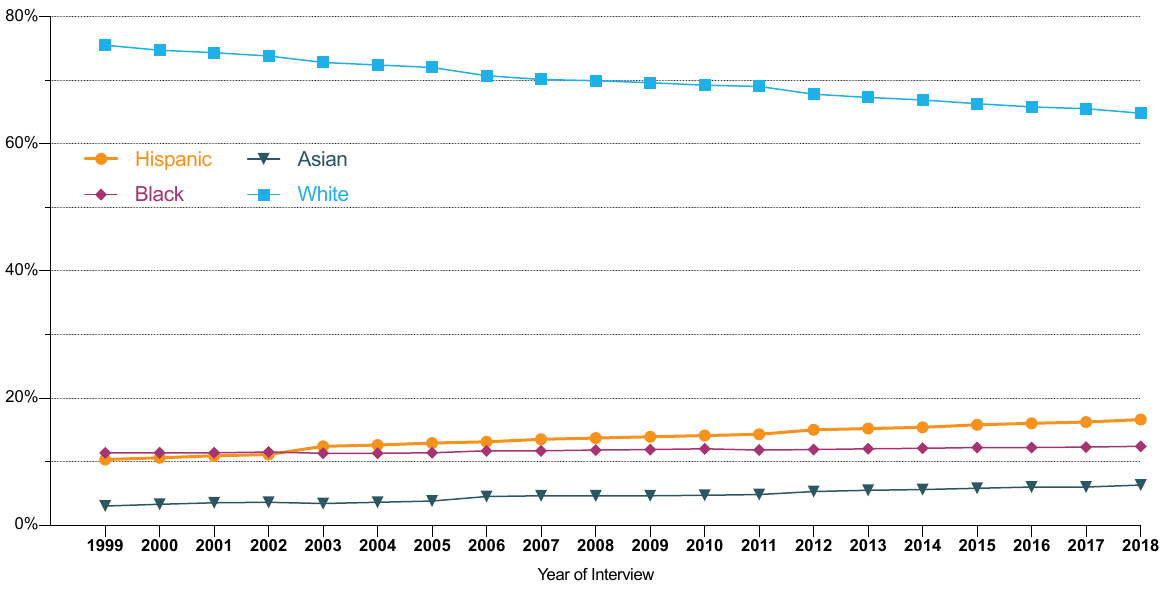
